# The use of home-based social care services in 2015/16 by individuals diagnosed with cancer in Scotland: a linked health and social care data analysis

**DOI:** 10.1101/2025.11.18.25340387

**Authors:** E Lemmon, D Henderson, S Clark, A Rutherford, CR Hanna, PS Hall

## Abstract

**Context:** Cancer is strongly associated with age. As detection and treatments improve, more individuals are living with and surviving the disease. Many of these individuals require social care as a direct result of their diagnosis and treatment effects, yet evidence on formal care receipt among people diagnosed with cancer is scarce.

**Objectives:** In this paper, we explore home-based, social care service use in Scotland among patients recently diagnosed with cancer and among those five-years post-diagnosis. We document differences between these two phases of care and explore variation between cancer types.

**Methods:** We conducted a retrospective cohort analysis of patients diagnosed with cancer in Scotland in 2015/16 and 2010/11. We used a unique dataset, combining – for the first time - cancer registrations data with Scotland’s Social Care Survey, to describe home-based social care use during the year 2015/16. We describe home-based social care use during 2015/16 in patients diagnosed with cancer in 2015/16 (Cohort 1) and 2010/11 (Cohort 2). We estimate multivariate regression models to explore differences between cancer types, controlling for demographic and clinical factors.

**Findings:** Among patients diagnosed with cancer in 2015/16, around 12% received some form of home-based social care during the same year compared with 10% of those diagnosed five years prior. We find significant differences in social care use depending on the type of cancer, even after controlling for age, sex, deprivation, geography and comorbidity.

**Limitations:** The research describes use of service only and cannot determine if this use reflects 0the needs of individuals or whether those needs are being met. Our five-year survivor cohort may be subject to survivorship bias which affects the interpretation of the results. Since we focus solely on those with a cancer diagnosis, we cannot conclude that use of home-based care is a direct result of a cancer diagnosis.

**Implications:** This work has shown that a substantial proportion of people diagnosed with cancer use home-based social care services. As the number of people living with and surviving cancer increases, understanding the evolving need for social care over time is necessary for future care planning and provision.

## 1. INTRODUCTION

In the UK, more than one-third of new cancer cases are in people aged 75 and over (Cancer Research UK, 2025a). It is estimated that by 2040, there will be a 20% increase in the number of new cases of cancer per year in the UK (Cancer Research UK, 2024). Survival has also continued to improve over time, thanks to screening programmes, new diagnostic techniques and advances in treatment. Around 50% of individuals diagnosed with cancer in 2018 will survive for 10 years or more, compared to just 24% in the 1970’s (Cancer Research UK, 2024).

Side effects from cancer and treatment in the immediate post-treatment period can leave patients with symptoms including neuropathy, pain, fatigue and mental health problems (Stein et al, 2008). At the same time, side effects can persist longer term and late effects can emerge years in the future. Late effects can include musculoskeletal complications, lymphedema, neurotoxicity, cardiac complications and more (Treanor et al, 2014). Moreover, cancer patients have poorer health outcomes compared to matched non-cancer patients (Yabroff et al, 2007).

The increasing incidence of cancer, an ageing population, and improving treatments, together with post-cancer symptoms create increased patient needs that may require long-term care (LTC) or social care providers. However, existing research in the area has noted the frequent myth that people with cancer do not have social care needs (MacMillan Cancer Support, 2015). Contrary to this belief, a MacMillan cancer support report demonstrated the widespread LTC needs of people living with and surviving cancer in the United Kingdom (UK) (MacMillan Cancer Support, 2015). The prevalence of cancer patients LTC needs has also been documented elsewhere (Boyes et al, 2012; Hodgkinson et al., 2007; Jacobs and Shulman, 2017; Nelson et al, 2015).

Understanding how individuals who are diagnosed with cancer use social care services, both at the time of treatment and perhaps more importantly as they survive the disease and continue to live into older ages, is crucial to ensure that those patients’ needs are met and for policy makers to adequately plan future service provision.

At present, much of the research into LTC use by individuals with cancer is focused on unpaid caregivers who provide essential support (Romito et al., 2013; Thomas et al, 2002).

We are aware of only two studies in the UK that have explored how individuals diagnosed with cancer use LTC services, such as those provided by local authorities and private agencies. The first study undertaken by MacMillan Cancer Research found that only 8% of individuals living with cancer who had practical and personal needs had those needs met by formal care services (MacMillan Cancer Support, 2015). Around 45% did not get any formal support at all and a further 11% got some formal support but did not feel it was enough (MacMillan Cancer Support, 2015). The second study, from the Nuffield Trust in 2014, looked at health and social care use among cancer patients in two localities in England, during the period around and following diagnosis and at the end of life (Chitnis X and M, 2014). Their research showed that around 10% of individuals diagnosed with cancer had a social care assessment within three months of being diagnosed. They also found that those diagnosed with cancer were less likely to use social care services in the year before their diagnosis and more likely to receive them in the 18 months after (Chitnis X and M, 2014). In their analysis of patients at the end of life, the study found that 42% of people with cancer were given a social care assessment. They also found that compared to people who died without cancer, 20% fewer individuals with cancer received social care in the last three months of life (Chitnis X and M, 2014).

Overall, the paucity of research exploring social care service use among people diagnosed with cancer has been recognised as a concern and can partly be attributed to the lack of available data (CRUK, 2025b). The unique collection of routine community social care data and advanced administrative data linkage infrastructure in Scotland can therefore contribute significantly to the evidence base. In this paper, we utilise Scotland’s 2015/16 Social Care Survey (SCS) (Henderson et al, 2019) and the cancer registration database (Public Health Scotland, 2025a) for individuals aged 50 and over. The linkage of these data sources allows us to report, for the first time, on the use of home-based social care services delivered to individuals living with cancer in Scotland. Specifically, we aim to address the following research questions:

- Does social care use differ between people recently diagnosed with cancer and those approximately five years post-diagnosis?
- How does social care use differ between cancer type?

## 2. DATA AND METHODS

### 2.1 Data

We utilise three key data sources from the project to describe the use of home-based social care services during the financial year 2015/16 among the Scottish population of patients aged 50 and over as of 1^st^ April 2015 and with a first diagnosis of cancer in either 2015/16 or 2010/11.

#### Demographics

The demographics file includes dates of birth, dates of death, sex, Scottish Index of Multiple Deprivation (SIMD) quintiles (Scottish Government, 2016), historic Charlson Comorbidity flags (from the period before 2010/11) including flags for an historic diagnosis of cancer or metastatic cancer (Charlson et al, 1987).

#### Social Care Survey

The Social Care Survey (SCS) is an historic, national collection of data on all local authority provided, adult community social care services delivered in Scotland, curated by the Scottish Government. This study uses SCS data for the financial year 2015/16 i.e., 1st April 2015 to 31^st^ March 2016. The SCS dataset captures delivery of local authority provided, home-based social care services including personal care (help with washing, dressing etc), home care including telecare, community alarms, meals services, help with laundry and shopping, self-directed support (SDS), housing support and social worker support. See Henderson et al (2019) for a full data resource profile.

#### Scottish Cancer Registry

The Scottish Cancer Registry (Scottish Morbidity Record 06 (SMR06)) contains records of all cancer diagnoses in Scotland (Public Health Scotland, 2025). We used data on all SMR06 registrations during the financial years 2010/11 and 2015/16.

#### Ethics and Information Governance

The project was approved by the Public Benefit Privacy Panel for Health and Social Care (HSC-PBPP) (project 1617-0012) and the linkage of the datasets was carried out by the Electronic Data Research and Innovation Service (eDRIS) Data and released to the research team in a pseudononymised format. All outputs were subject to rigorous disclosure control checks before being released.

### 2.2 Methods

We carry out a retrospective, individual level analysis to describe the use of home-based social care services during the financial year 2015/16 in patients aged 50 and over diagnosed with a first cancer in 2015/16 (Cohort 1) and 2010/11 (Cohort 2).

#### Cohort selection

The initial population is made up of 214,505 individuals with a single diagnosis of cancer recorded in the cancer registry during the period 2010/11 to 2015/16 and still alive on 30^th^ March 2016. After dropping individuals with a cancer diagnosis in the period before 2010/11 (n = 78,762) or during 2012/13-2014/15 (n = 88,081); those with an in-situ cancer (n = 4,844); those with non-melanoma skin cancers (International Disease Classification 10 code C44) (World Health Organisation, 2019) (n = 13,345), the final cohort is made up of N = 29,473 unique individuals. From here, we created two distinct cohorts to examine social care use at two clinically relevant time points following diagnosis. This approach was necessary since the SCS data were available for financial year 2015/16 only. Thus, the two cohorts - Cohort 1 (diagnosed 2015/16) (N_1_ = 18,729) and Cohort 2 (diagnosed 2010/11) (N_2_ = 10,744)- represent two phases in the cancer survivorship pathway, rather than longitudinal follow-up of the same individuals.

We present descriptive characteristics of the two cohorts and describe their use of social care services in 2015/16. We also estimate a set of multivariate regression models to explore differences in use of social care between the most common cancer diagnoses, accounting for common demographic characteristics. Specifically, regression outcomes are: receipt of any home-based social care service; receipt of personal care among those who received any type of care; and hours of personal care received among those who received personal care. Demographic information includes sex, age, urban rural six-fold classification (Scottish Government, 2022), Scottish Index of Multiple Deprivation (SIMD) quintile (Scottish Government, 2020) and number of Charlson comorbidities (Charlson et al., 1987).

## 3. RESULTS

Table 1 displays the descriptive statistics for the two cohorts. The final rows of the table show that 11.54% (n = 2,161) of individuals diagnosed with cancer in 2015/16 received some form of social care service during 2015/16. This is compared to 10.41% (n = 1,118) of those diagnosed in 2010/11 and this difference is significant at the 1% level. In terms of patient characteristics, Cohort 2 (diagnosed in 2010/11) were significantly more likely to be female, older, less deprived, less urban and more comorbid. Most likely these trends reflect the cohort selection requiring individuals to be alive at the end of 2015/16 (in order to observe their social care use) meaning that people with poorer prognoses for cancers diagnosed in 2010/11, are less likely to survive up until that point.

**Table 1.**
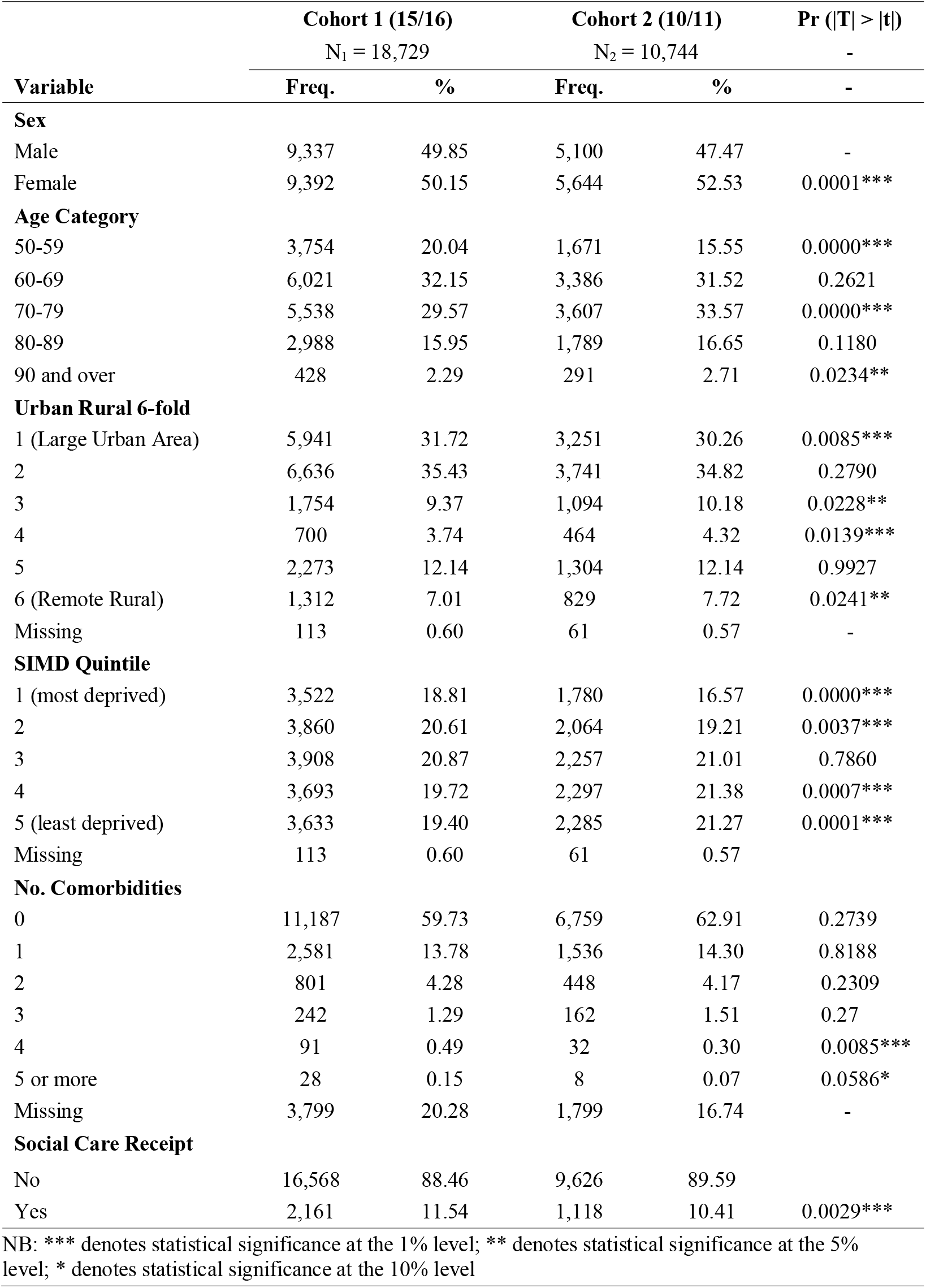
Patient characteristics by diagnosis year.

Table 2 presents the most common cancer types across both cohorts. In both cohorts, and in line with published national statistics, breast and prostate cancers were the most common cancers. The distribution of bronchus and lung cancer patients differs substantially between the two cohorts – 11.56% and 2.81% for Cohort 1 and 2 respectively - most likely reflecting the poorer survival rates associated with lung cancer meaning that less patients with a diagnosis were still alive in 2015/16 and therefore present in Cohort 2.

**Table 2.**
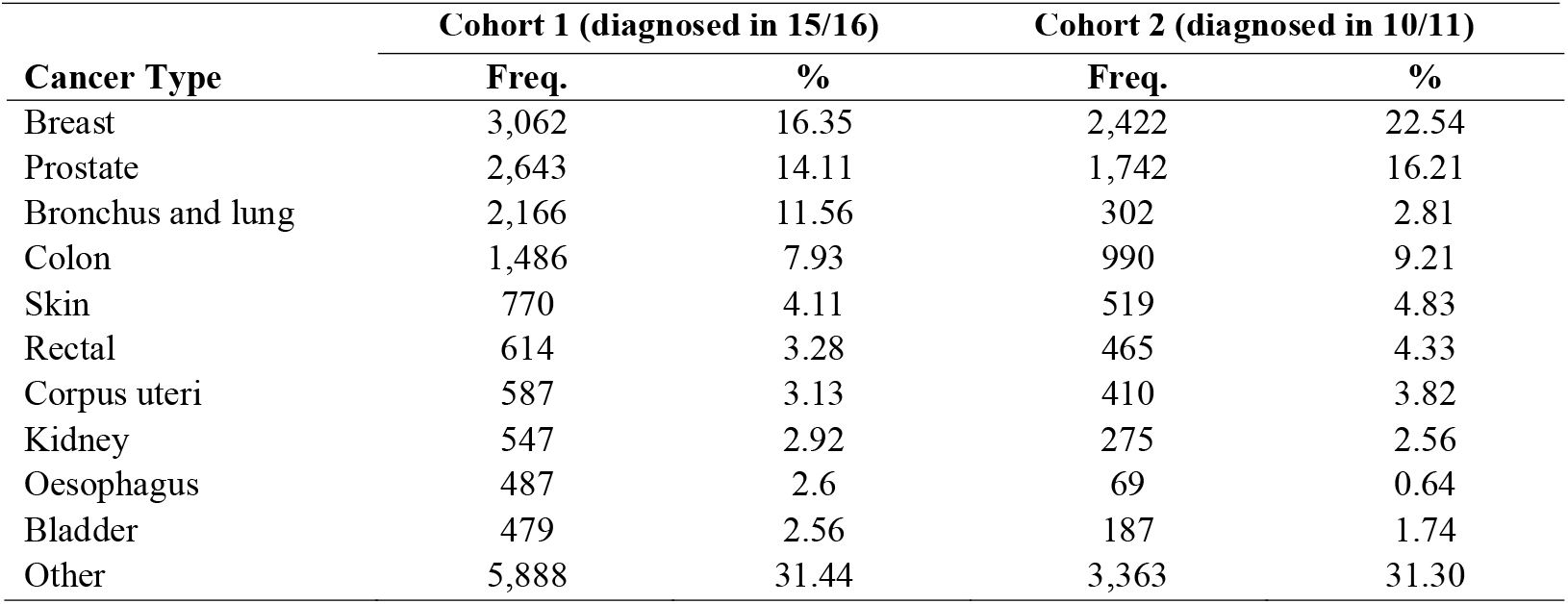
Ten most commonly diagnosed cancers across both cohorts.

Fig. 1 shows the proportion of individuals receiving some form of social care service within the ten most commonly diagnosed cancers across both cohorts. Among those individuals diagnosed during 2015/16, individuals diagnosed with lung cancer were most likely to receive some form of social care (18.1%). This is compared to 14.6% of lung cancer patients diagnosed in 2010/11. The highest rate of social care use is observed among those diagnosed with bladder cancer in 2010/11. Specifically, over 20% of patients diagnosed with bladder cancer in 2010/11 received social care in 2015/16. This is compared to just 12% of patients diagnosed with bladder cancer during the same year. A similar pattern can be seen for patients diagnosed with oesophageal cancer, where a larger proportion of patients diagnosed in 2010/11 (14.5%) received care compared to those diagnosed in 2015/16 (10.1%). A sizeable difference in care receipt is also apparent among patients diagnosed with rectal cancer. In particular, for patients diagnosed during 2015/16, around 11.4% received some form of care, compared to 6.7% diagnosed five years previously.

**Figure 1.**
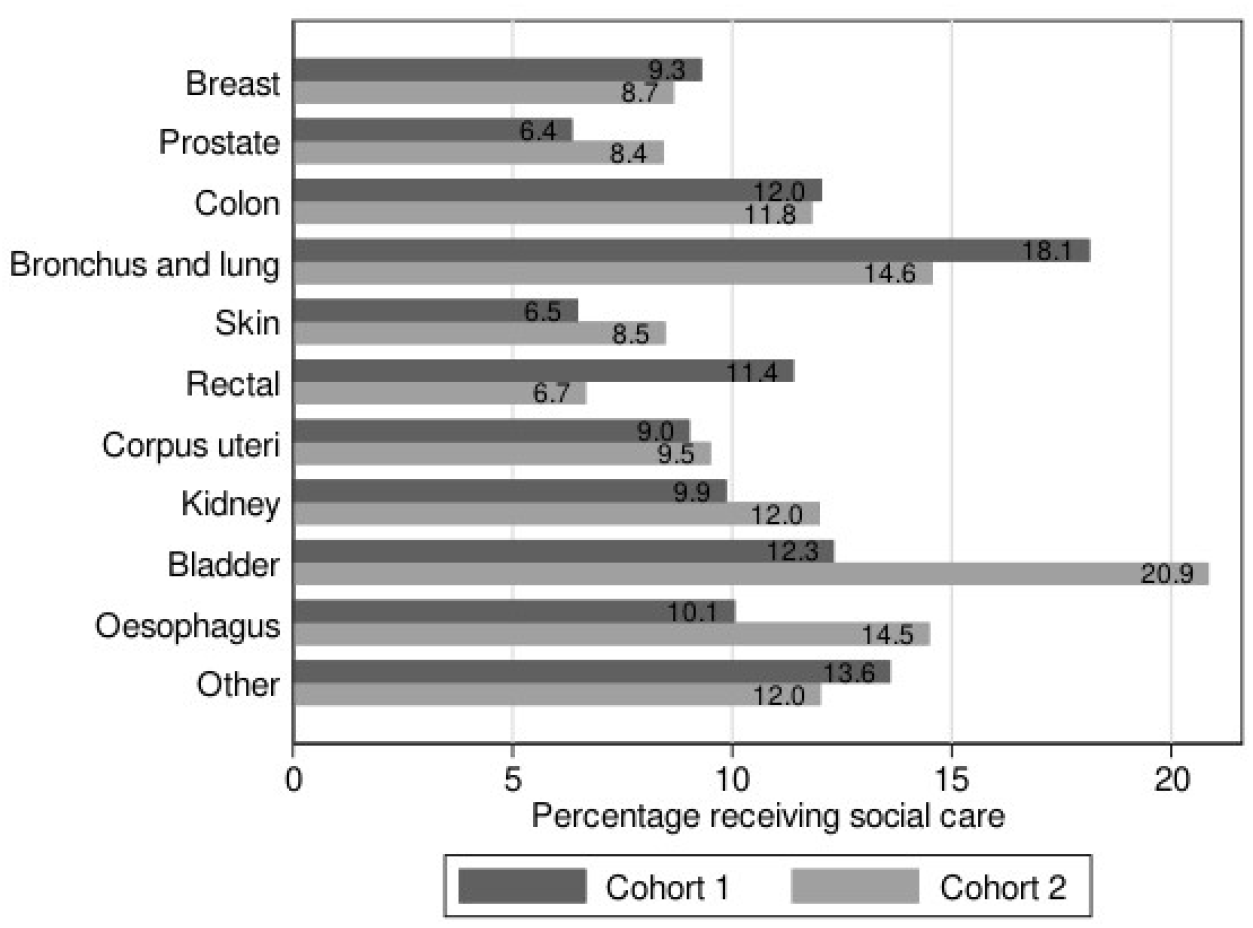
Percentage of patients receiving social care in 2015/16 within the ten most common cancers in Cohort 1 (diagnosed 2015/16) and Cohort 2 (diagnosed 2010/11)

In Table 3, type of social care service provision is shown for those individuals who received social care within each cohort. Those services are home care, personal care, meals services, self-directed support, housing support, telealarm and other telecare services. Mean personal care hours are also shown for those individuals who received personal care services. Overall, the type of services received in the two cohorts are similar but those diagnosed in 2010/11 tended to have a higher use of telecare, community alarm, meals and housing support services, though these differences were not statistically significant. The mean hours of personal care received in the 2015/16 cohort were 8.4 hours per week compared to 7.8 hours in the 2010/11 cohort. This difference was statistically significant at the 10% level. Those diagnosed in 2015/16 had a slightly higher receipt of home care services (43% compared to 41%), but this difference was not statistically significant.

**Table 3.** Type of social care services among patients who received social care in 15/16.

| Variable | Cohort 1 (15/16) |  | Cohort 2 (10/11) |  | Pr(\|T\| > \|t\|) |
| --- | --- | --- | --- | --- | --- |
|  | n = 2,161 |  | n = 1,118 |  |  |
|  | Freq. | % | Freq. | % |  |
| **Home Care** |  |  |  |  |  |
| 0 | 1,234 | 57.10 | 665 | 59.48 | - |
| 1 | 927 | 42.90 | 453 | 40.52 | 0.1912 |
| **Personal Care** |  |  |  |  |  |
| 0 | 1,294 | 59.88 | 681 | 60.91 | - |
| 1 | 867 | 40.12 | 437 | 39.09 | 0.5670 |
| Mean personal care hours | 8.4 | - | 7.8 | - | 0.0778* |
| **Meals** |  |  |  |  |  |
| 0 | 2,068 | 95.70 | 1,067 | 95.44 | - |
| 1 | 93 | 4.30 | 51 | 4.56 | 0.7325 |
| **SDS** |  |  |  |  |  |
| 0 | 1,645 | 76.12 | 852 | 76.21 | - |
| 1 | 516 | 23.88 | 266 | 23.79 | 0.9567 |
| **Housing Support** |  |  |  |  |  |
| 0 | 2,005 | 92.78 | 1,021 | 91.32 | - |
| 1 | 156 | 7.22 | 97 | 8.68 | 0.1383 |
| **Alarm** |  |  |  |  |  |
| 0 | 676 | 31.28 | 317 | 28.35 | - |
| 1 | 1,485 | 68.72 | 801 | 71.65 | 0.0838* |
| **Telecare** |  |  |  |  |  |
| 0 | 1,761 | 81.49 | 892 | 79.79 | - |
| 1 | 400 | 18.51 | 226 | 20.21 | 0.2392 |

NB: \*\*\* denotes statistical significance at the 1% level; \*\* denotes statistical significance at the 5% level; \* denotes statistical significance at the 10% level

Fig. 2 shows mean hours of weekly personal care received for the top ten most commonly diagnosed cancers across the two groups. For individuals diagnosed in 2015/16 average weekly hours of personal care ranged from 11 hours per week for those diagnosed with kidney cancer and 6.5 hours per week for those diagnosed with bladder cancer. In the cohort of individuals diagnosed five years prior, average hours of personal care were highest for breast cancer patients at 9.2 hours per week, and lowest for prostate cancer patients at 6.6 hours per week.

**Figure 2.**
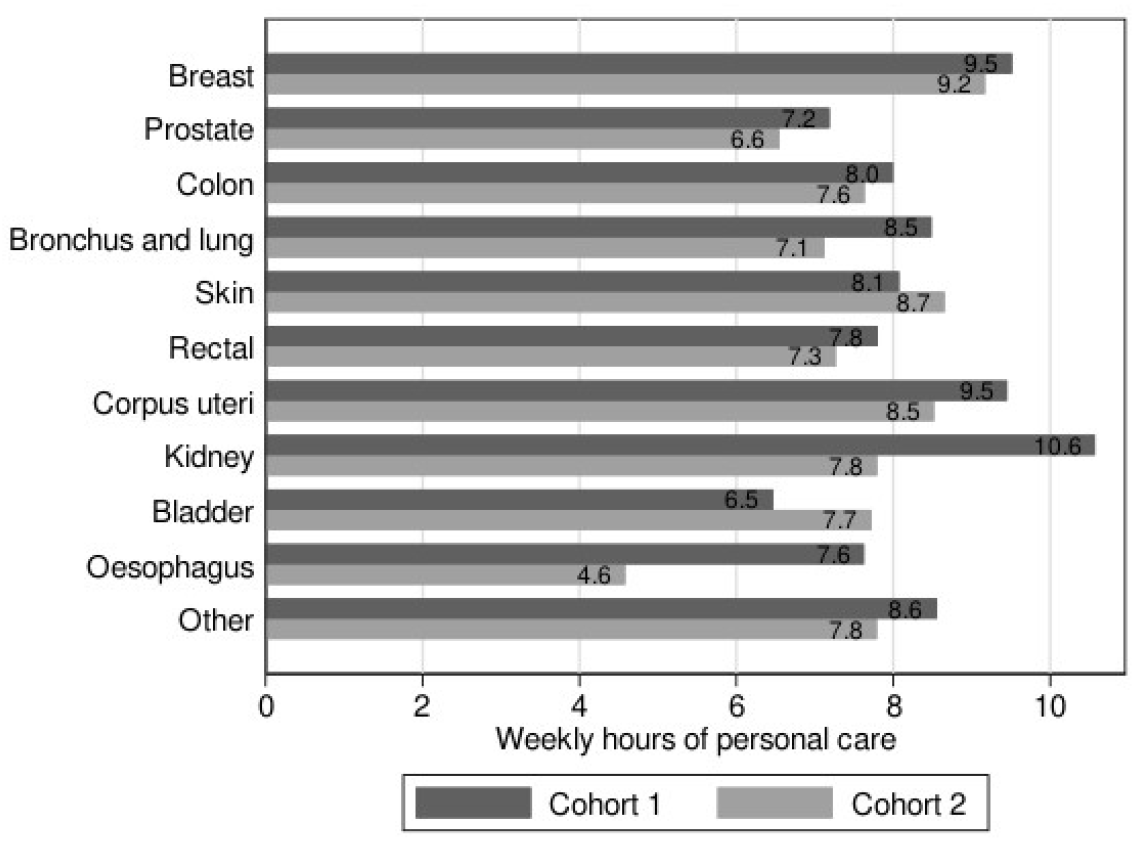
Mean hours of personal care received in Cohort 1 (diagnosed 2015/16) and Cohort 2 (diagnosed 2010/11)

Finally, Table 4 and Table 5 display the regression results from three regressions for each cohort. The first is a probit model for the likelihood of receipt of any social care service. The second is a probit model for the likelihood of receipt of personal care, among those individuals who received some form of social care. The last is an Ordinary Least Squares (OLS) regression of weekly hours of personal care amongst those individuals who received personal care. Coefficients and p-values are shown, with statistical significance indicated by asterisks. All models use robust standard errors and control for sex, age category, urban-rural six-fold indicator, SIMD quintile and number of comorbidities.

**Table 4.**
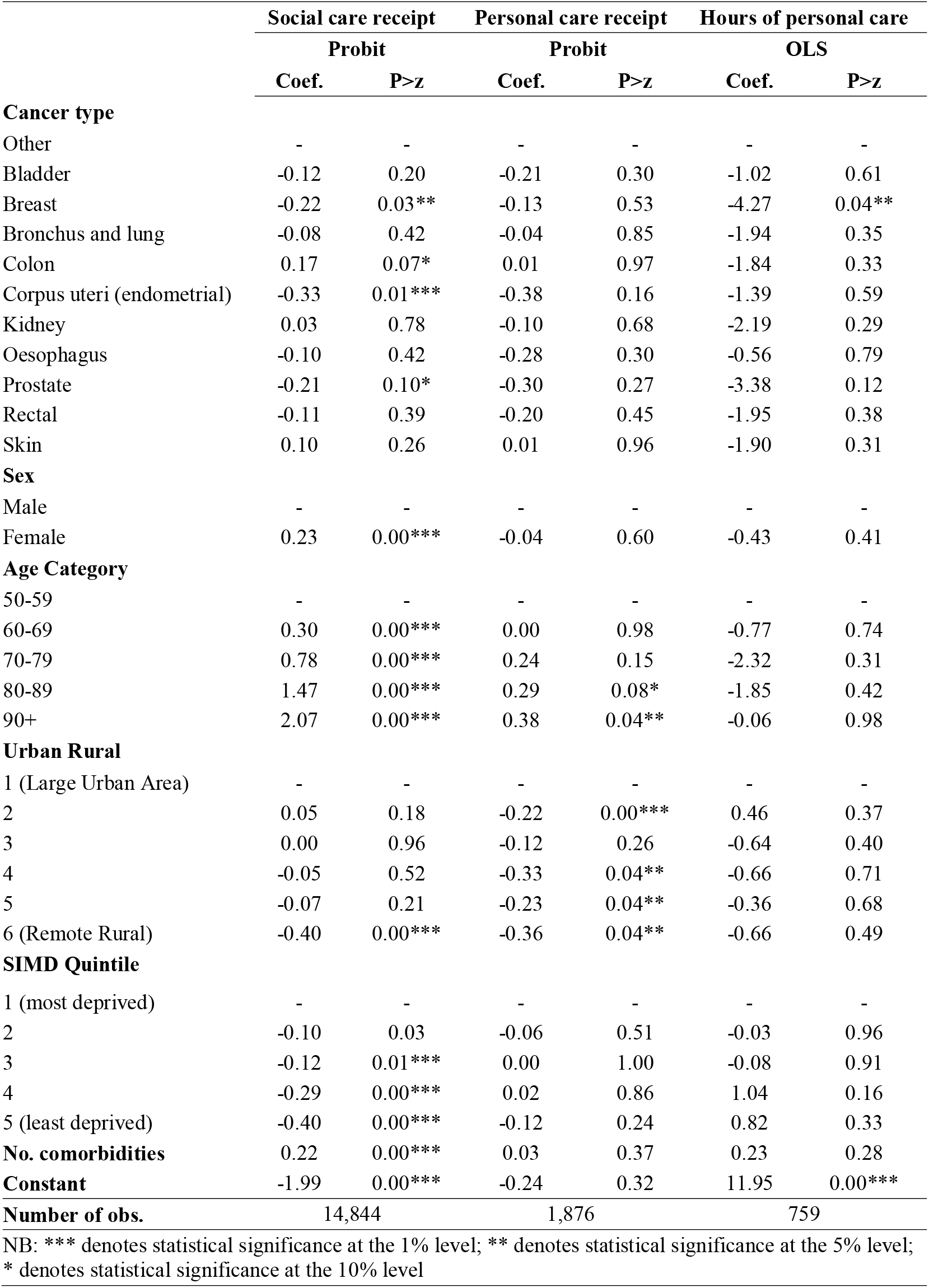
Regressions for Cohort 1 (diagnosed 15/16)

**Table 5.** Regressions for Cohort 2 (diagnosed 10/11)

|  | Social care receipt |  | Personal care receipt |  | Hours of personal care |  |
| --- | --- | --- | --- | --- | --- | --- |
|  | Probit |  | Probit |  | OLS |  |
|  | Coef. | P>z | Coef. | P>z | Coef. | P>z |
| **Cancer type** |  |  |  |  |  |  |
| Other | - | - | - | - | - | - |
| Bladder | -0.33 | 0.01*** | 0.19 | 0.48 | -0.64 | 0.79 |
| Breast | -0.27 | 0.04** | 0.08 | 0.77 | -1.77 | 0.47 |
| Bronchus and lung | -0.29 | 0.03** | 0.06 | 0.84 | -1.92 | 0.42 |
| Colon | -0.16 | 0.29 | 0.43 | 0.18 | -2.47 | 0.31 |
| Corpus uteri (endometrial) | -0.41 | 0.01*** | 0.08 | 0.81 | -0.86 | 0.74 |
| Kidney | -0.54 | 0.00*** | 0.64 | 0.08 | -2.74 | 0.29 |
| Oesophagus | -0.26 | 0.10* | 0.19 | 0.58 | -0.18 | 0.95 |
| Prostate | 0.09 | 0.60 | 0.07 | 0.84 | -1.07 | 0.70 |
| Rectal | -0.14 | 0.54 | 0.26 | 0.58 | -3.92 | 0.13 |
| Skin | -0.15 | 0.21 | 0.23 | 0.37 | -1.82 | 0.42 |
| **Sex** |  |  |  |  |  |  |
| Male | - | - | - | - | - | - |
| Female | 0.31 | 0.00*** | -0.03 | 0.79 | 0.90 | 0.21 |
| **Age Category** |  |  |  |  |  |  |
| 50-59 | - | - | - | - | - | - |
| 60-69 | 0.37 | 0.00*** | 0.55 | 0.06* | -5.22 | 0.29 |
| 70-79 | 0.66 | 0.00*** | 0.73 | 0.01*** | -5.39 | 0.27 |
| 80-89 | 1.50 | 0.00*** | 0.77 | 0.01*** | -5.59 | 0.25 |
| 90+ | 2.00 | 0.00*** | 0.90 | 0.00*** | -4.24 | 0.39 |
| **Urban Rural** |  |  |  |  |  |  |
| 1 (Large Urban Area) | - | - | - | - | - | - |
| 2 | 0.11 | 0.02** | -0.11 | 0.29 | 0.68 | 0.31 |
| 3 | 0.05 | 0.46 | 0.03 | 0.85 | 1.40 | 0.25 |
| 4 | 0.05 | 0.60 | 0.19 | 0.36 | 1.46 | 0.22 |
| 5 | 0.00 | 1.00 | -0.06 | 0.70 | 1.60 | 0.22 |
| 6 (Remote Rural) | 0.00 | 0.96 | -0.06 | 0.73 | 0.26 | 0.83 |
| **SIMD Quintile** |  |  |  |  |  |  |
| 1 (Most deprived) | - | - | - | - | - | - |
| 2 | -0.14 | 0.03** | -0.04 | 0.76 | -1.25 | 0.11 |
| 3 | -0.07 | 0.26 | -0.20 | 0.13 | -1.34 | 0.10* |
| 4 | -0.30 | 0.00*** | -0.12 | 0.38 | 0.32 | 0.78 |
| 5 (Least Deprived) | -0.37 | 0.00*** | -0.39 | 0.01*** | -0.13 | 0.89 |
| **No. comorbidities** | 0.21 | 0.00*** | 0.09 | 0.04** | 0.55 | 0.06* |
| **Constant** | -1.95 | 0.00*** | -1.05 | 0.01*** | 13.47 | 0.01*** |
| **Number of obs.** | 8,894 |  | 998 |  | 392 |  |

NB: \*\*\* denotes statistical significance at the 1% level; \*\* denotes statistical significance at the 5% level; \* denotes statistical significance at the 10% level

In both cohorts, females are significantly more likely to receive social care compared to males. The likelihood of care receipt also increases with age and the number of comorbidities in both cohorts. Moreover, receipt of social care is less likely with lower levels of deprivation. In those patients diagnosed in 2015/16, living in the most rural settings is associated with a significantly higher likelihood of social care service use compared to the most urban setting. Whereas, in those diagnosed in 2010/11, living in the second most urban area is associated with a higher likelihood of social care service use compared to the most urban setting. The probit models for social care receipt in both cohorts demonstrate some statistically significant differences between cancer types, even after accounting for relevant demographic and clinical factors.

## CONCLUSION AND DISCUSSION

In this paper, we set out to shed light on an important issue for cancer care where current evidence is lacking. Specifically, we aimed to understand if social care service use differs between people recently diagnosed with cancer and those approximately five-years post diagnosis. Further, we sought to determine whether there are differences in social care service use between cancer types.

### Does social care service use differ between people recently diagnosed with cancer and those approximately five-years post diagnosis?

Our findings show that among individuals diagnosed with cancer in 2015/16, around 11.54% used some form of social care during the year of diagnosis. This is compared to 10.41% for individuals approximately five years post-diagnosis (diagnosed in 2010/11). Whilst this difference was statistically significant at the 1% level (Table 1), the absolute difference is relatively small, implying that social care needs persist among cancer survivors in the years following diagnosis. These figures also align with the limited published evidence on social care service use amongst cancer patients. Specifically, evidence from MacMillan’s 2015 report suggested 10% of patients received some form of social care service MacMillan Cancer Support (2015). Their cohort included people in the immediate period of diagnosis right up until end of life. It is also important to note that Cohort 2 (diagnosed in 2010/11) contains only individuals who survived at least five years post-diagnosis and thus will exclude individuals with poor prognosis cancer such as lung and oesophageal. As a result, the lower rate of observed social care use amongst this cohort compared to those recently diagnosed, may reflect survivor bias rather than any reduced level of need over time.

Amongst those individuals who received some form of social care, the most common type of care received in both cohorts was a community alarm. A higher proportion of individuals diagnosed in 2010/11 had a community alarm compared to those diagnosed in 2015/16 (72% and 69% respectively), a difference that was statistically significant at the 10% level (Table 3). Mean personal care hours were statistically significantly lower among the five-year survivor cohort (7.8 hours per week) compared with those diagnosed in 2015/16 (8.4 hours). Once again, although these differences are statistically significant, they may reflect survivorship bias since the five-year survivor cohort contains relatively more healthy individuals. For other types of care e.g., personal care, meals, SDS, telecare and housing support, there was no difference in care use between the two groups.

When we look at care receipt within cancer types, there appear to be more pronounced differences between the two phases of care, possibly offering more insight into changing care needs over the cancer survivorship pathway. For example, among patients diagnosed with bladder or oesophageal cancer, a higher proportion of those diagnosed in 2010/11 received care in 2015/16 compared to those diagnosed during 2015/16 (Fig.1). This could reflect an increasing requirement for support in the years post diagnosis for these cancer types, however it may also reflect the fact that only patients who survived to 2015/16, and who may have ongoing morbidity, are included in the cohort. In contrast, the opposite is true for patients diagnosed with bronchus and lung, and rectal cancer, where a higher proportion of individuals received care when diagnosed during the same year i.e., 2015/16. For these poorer prognosis cancers, this pattern may reflect high levels of need around the time of diagnosis combined with substantial mortality before five-year survival. For other cancer types, for example breast cancer, the proportion receiving care appears relatively similar across the two phases, suggesting more stable patterns of support over time.

In terms of intensity of support received, mean hours of personal care tended to be higher among individuals diagnosed in 2015/16 compared to those diagnosed five years prior, with the exception of bladder and skin cancers (Fig. 2). This could reflect a reduced need for care in the years post diagnosis, however the interpretation is once again complicated by survivorship bias in the survivor cohort, where individuals with advanced disease, high levels of disability and significant comorbidity are underrepresented. For patients diagnosed with skin and bladder cancer, the opposite is true. Specifically, mean hours of care were higher among those diagnosed in 2010/11 compared to in 2015/16. These cancers have relatively higher survival rates, and persistent or late-emerging morbidity may explain the higher personal care hours observed among five-year survivors.

### How does social care service use differ between cancer types?

The descriptive statistics suggest some differences in the use of social care services between cancer types. Fig. 1 demonstrates that amongst the 10 most common cancers diagnosed across the two cohorts, for those individuals diagnosed in 2015/16, there was an 11.7 percentage point difference in care receipt between patients diagnosed with lung cancer (18.1% received social care) and patients diagnosed with prostate cancer (6.4% received social care). For those diagnosed in 2010/11, the biggest difference in care receipt was of 14.2 percentage points, this time between individuals diagnosed with bladder cancer (20.9% in receipt of care) and individuals diagnosed with rectal cancer (6.7% in receipt of care). Fig. 2 shows that there are also visible differences in personal care intensity between cancers. The largest difference among individuals diagnosed in 2015/16 being between kidney cancer (10.6 hours per week) and bladder cancer (6.5 hours per week). Whereas for those diagnosed in 2010/11, the biggest difference in hours of personal care received per week was between breast cancer (9.2 hours per week) and oesophageal cancer (4.6 hours per week). These differences may reflect differences in the need for care by cancer type. For example, due to symptoms associated with a specific cancer and its treatment or due to patient characteristics associated with particular cancers.

The regression models attempt to control for some of these other factors, including age, sex, deprivation, geography and level of morbidity, and still indicate some significant differences in the predicted probability of care receipt. For individuals diagnosed in 2015/16, compared to the least common types of cancer, patients diagnosed with breast, endometrial and prostate cancer were significantly less likely to receive any form of social care. Whereas individuals diagnosed with colon cancer, are significantly more likely to receive care. These differences disappear when looking at personal care receipt. For personal care intensity, the only significant difference in personal care hours received is between those diagnosed with breast cancer in 2015/16, who received significantly less personal care hours compared to the least common cancer types. Among those diagnosed in 2010/11, compared to patients diagnosed with the least common types of cancer during the same period, individuals diagnosed with bladder, breast, lung, endometrial, kidney and oesophageal cancer, were significantly less likely to receive any form of social care in 2015/16. Once again suggesting that even after accounting for other relevant factors, care receipt differs between cancer types.

Whilst these results are an important step forward in shedding light on a previously under-researched area, there are some limitations to consider. Firstly, the underlying data demonstrate care receipt only. They do not indicate the extent of need among the cancer population nor reflect the extent to which needs are being met. Related to this, we have not considered varying levels of need that is dependent on cancer type which would help to determine if the observed differences in service use are indeed a reflection of the needs associated with specific cancers. A second limitation of this study, as we have discussed when interpreting the results, is the potential survivorship bias among the five-year survivor cohort. Since we have conditioned this cohort on surviving up to 2015/16, it necessarily will exclude poorer prognosis cancers and higher needs. This means that the 2010/11 cohort may underrepresent individuals with high needs. A further limitation is that we have restricted it to contain only those with a cancer diagnosis. As such, we offer no comparison to a non-cancer population which would allow us to get closer to determining the causal impact of a cancer diagnosis on the use of home-based social care services in Scotland. Thirdly, there may be other relevant factors that we have not accounted for, which may determine social care use among the cancer population. Whilst we have attempted to account for comorbidities, this comes from hospitalisations records so is likely to be an underestimate of the level of comorbidity in the population. There may be additional needs-based indicators that would capture the level of need in a cohort. Not controlling for these confounders may impact the results. Finally, the data are somewhat out of date and we would suggest that future research capitalises on the new version of the SCS, Source, which is now curated by Public Health Scotland (Public Health Scotland, 2025b).

Notwithstanding these limitations, this paper has, for the first time, explored use of home-based social care services amongst the population of individuals diagnosed with cancer in Scotland, by linking Scottish cancer registrations data to the SCS. The results have shown that a substantial proportion of people diagnosed with cancer rely on social care services, both at the time of diagnosis and five years post diagnosis. Finally, the results have pointed to significant differences in social care use depending on the type of cancer, even after controlling for other relevant confounding factors. Further research is warranted to understand whether this use is aligned with patient need and whether or not a cancer diagnosis has a direct impact on home-based social care service use.

## Data Availability

Data are not publicly available.

## Acknowledgements and Declarations

Approval to access this data was granted by the Scottish Government in January 2017. Project number: SG000-000850. This work uses data provided by patients and clients, collected by the NHS and the Scottish Government as part of their care and support. The authors would like to acknowledge the eDRIS team (Public Health Scotland) for their support in obtaining approvals, the provisioning and linking of data and facilitating access to the National Safe Haven.

